# Analysis of clinical characteristics of invasive non-typhoidal *Salmonella* infections in children: a 5-year dual-center retrospective study

**DOI:** 10.1101/2024.10.03.24314827

**Authors:** Chenggang Lu, Yongping Xie, Yan Li, Fangfang Cheng, Lisu Huang, Wang Hua

## Abstract

**Objective:** To investigate the incidence, microbiological characteristics, clinical manifestations, and antibiotic resistance of invasive non-typhoidal *Salmonella* (iNTS) infections in children.

**Methods:** A retrospective study was conducted from January 2018 to December 2022 at two large teaching hospitals: Zhejiang University School of Medicine Children’s Hospital (ZCH) and Children’s Hospital of Soochow University (SCH). Medical records of culture-confirmed iNTS cases were reviewed, and a standardized case report form was used to collect demographic, clinical, and microbiological data.

**Results:** A total of 109 cases of iNTS infection were included, with 40 cases from SCH and 69 cases from ZCH. 71 cases (65.1%) were boys.. Infants under 1 year old accounted for 30.3% of the cases. Among the cases, 38 cases (34.9%) had underlying diseases, mainly tumors (55.3%,21/38). The most common sites of infection were the bloodstream (84.4%) and bones and joints (12.8%). The most common serogroups were B (36.8%), C (44.7%) and D (38.2%). The overall resistance rate of ampicillin was 53.6%, ceftriaxone resistance rate was 17.8%, and carbapenem resistance rate was 0%. Seasonal trends indicated higher incidence rates during the summer.

**Conclusion:** iNTS infections in children primarily affect the bloodstream and bones/joints, with a significant proportion of cases in those with underlying medical conditions. Ceftriaxone remains the first-line antibiotic, but increasing resistance highlights the need for vigilant antimicrobial stewardship. Carbapenems can be considered as second-line antibiotics in severe cases. Further studies are needed to understand the molecular characteristics and improve treatment strategies for iNTS infections in pediatric patients.

## Introduction

*Salmonella* is easily transmitted to the human body through foodborne and direct contact routes, and it is a common pathogen in pediatric gastrointestinal infections. Some cases present with extraintestinal infections, including bacteremia and secondary focal lesions. A systematic review estimated(1)that in 2017, there were 535,000 cases of invasive non-typhoidal *salmonella* (NTS) disease globally, resulting in 77,500 deaths. Megged et. al report that the age characteristics of NTS bloodstream infection show a U-shaped distribution and were more common in children and the elderly(2). The highest incidence rate of invasive non-typhoidal *salmonella* disease was observed in children under the age of 5, reaching 34.3 per 100,000 person-years(1). According to a systematic review and analysis of literature on NTS invasive disease cases before June 2021, the overall pooled case-fatality ratio estimates were as follows: 14.7% in Africa, 14.0% in Asia, 9.6% in the Americas, and 9.9% in Europe(3). Data from the provinces of Zhejiang and Henan in China indicate that 20.9% to 43.6% of NTS cases result in invasive infections, with 52.8% to 67.9% of these cases being bloodstream infections. Furthermore, more than half of the cases affected children(4). Bloodstream infections without appropriate antibiotic treatment are fatal, and changes in antibiotic resistance spectrum can lead to treatment failure and dissemination of infection. To date, there is a lack of comprehensive and in-depth understanding among pediatric clinicians regarding invasive salmonella infections, becoming a clinically significant issue that cannot be ignored.

## Materials and Methods

### Study Population and Data Collection

The study included all subtypes of NTS except for typhoid and paratyphoid strains. Invasive non-typhoidal *Salmonella* infections (iNTS) were defined as cases where specimens cultured from any normally sterile site were positive. The serogroup of the Salmonella isolates was determined using slide agglutination with antisera, and the isolates were classified into seven serogroups: serogroup A-E and other serogroups. Antimicrobial susceptibility testing was determined using a commercial microdilution method (VITEK COMPACT, BioMerieux, France), and the results were interpreted according to Clinical Laboratory Standards Institute guideline M100-ED32. Susceptibility to ampicillin, ceftriaxone, chloramphenicol, imipenem, ciprofloxacin, and trimethoprim/sulfamethoxazole (SMZ) was tested. E. coli ATCC25922 was used for quality control. A retrospective analysis was conducted on iNTS data from the Zhejiang University School of Medicine Children’s Hospital (ZCH) and the Children’s Hospital of Soochow University (SCH) from January 2018 to December 2022. Medical records of confirmed cases were retrieved, and a standardized case report form was used to collect information on age, gender, underlying medical history, symptoms, laboratory examinations, and antimicrobial susceptibility. The pediatric sequential organ failure assessment (pSOFA) score was evaluated according to the previous literature(5).Meteorological data were obtained from the Zhejiang Government Services Network (www.zjzwfw.gov.cn) and JiangSu Meteorological Administration (js.cma.gov.cn).

### Statistical Analysis

Non-normal variables were presented as median and interquartile range; and categorical variables were described as frequencies.Differences in continuous variables between the bloodstream infection (BSI) group and non-BSI group, as well as between the age >1 year group and the age ≤ 1 year group, were compared using the nonparametric Mann-Whitney U test. Proportions were compared using the χ-squared test. *P* <0.05 was considered statistically significant. All statistical analyses were performed using SPSS statistical 27.0 software (IBM, Armonk, NY, USA).

### Ethics statement

The study was approved by Medical Ethics Committee of the Zhejiang University School of Medicine Children’s Hospital and the Children’s Hospital of Soochow University (2024-IRB-0193-P-01). Written assent forms were obtained from all participants, along with written consent forms from their guardians.

## Results

109 cases of iNTS infection were included in the study, with 40 cases from SCH and 69 cases from ZCH. Of these, 71 cases (65.1%) were boys. Additionally, there were 33 infants (30.3%) under one year old. 38 cases (34.9%) had underlying diseases, mainly tumors (21 cases), including 19 cases of acute leukemia, 1 case of bone tumor, and 1 case of optic nerve neuroepithelial tumor. None of the cases were co-infected with HIV. There were 92 cases (84.4%) of bloodstream infection, 7 cases of bone marrow infection, 7 cases of joint infection, 2 cases of central nervous system infection, and 1 case of pleural effusion. The median time to positive blood culture was 15.8 hours, and the median length of hospital stay was 12 days.There were three deaths, all from ZCH, two of which were boys with acute lymphoblastic leukemia and *Salmonella* group D infection, and one was a girl with acute monocytic leukemia and *Salmonella* group B infection. They had significantly elevated pSOFA scores. All three cases had decreased platelet counts and significantly elevated CRP levels.

In terms of monthly case numbers and temperature, the curve trends suggest a clear correlation between temperature and the number of cases in both children’s hospitals, which increasing markedly in summer when temperatures are high. while the correlation with relative humidity is not obvious. (Fig.1)

**Fig. 1.**
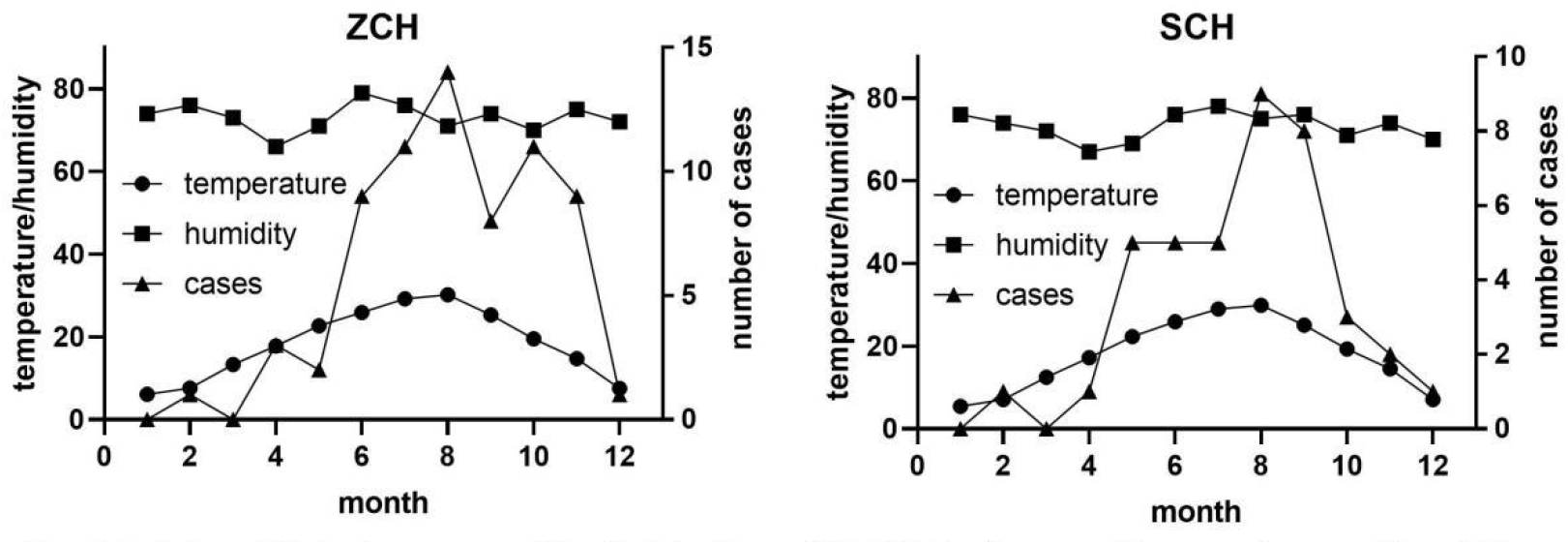
Relationship between monthly distribution of iNTS infections and temperature and humidity.

Compared to the BSI group, the non-BSI group had a significantly higher incidence of bone and joint symptoms (52.9% vs. 4.3%, *P* < 0.001), higher white blood cell count[7.9×10^9^/L vs. 11.1×10^9^/L, *P*=0.009], higher platelet count [226.5×10^9^/L vs. 414.0×10^9^/L, *P* < 0.001], and longer duration of effective antibiotic use (9.0days vs. 25.0days, *P* < 0.001). There were no significant differences in other indicators. (Tables 1)

**Table 1.** Demographic and clinical manifestations of children with iNTS infections, 2018–2022.

| Variables | n | BSI | n | Non-BSI iNTS | $\chi^2/Z$ | *P* | n | Age >1 year | n | Age ≤ 1 year | $\chi^2/Z$ | *P* |
| --- | --- | --- | --- | --- | --- | --- | --- | --- | --- | --- | --- | --- |
| Demographic |  |  |  |  |  |  |  |  |  |  |  |  |
| Boys, n (%) | 92 | 59(64.1) | 17 | 12(70.6) | 0.264 | 0.608 | 76 | 50(65.8) | 33 | 21(63.6) | 0.047 | 0.828 |
| Age, months, median (IQR) | 92 | 19.5(12.0,56.3) | 17 | 17.0(10.1,23.0) | -1.374 | 0.169 | / | / | / | / | / | / |
| Underlying diseases, n (%) | 92 | 34(37.0) | 17 | 4(23.5) | 1.139 | 0.286 | 76 | 31(40.8) | 33 | 7(21.2) | 3.883 | 0.049 |
| Tumors, n (%) | 92 | 20(21.7) | 17 | 1(5.9) | 1.412 | 0.235 | 76 | 21(27.6) | 33 | 0(0) | 11.294 | 0.001 |
| Deep vein catheter, n (%) | 92 | 23(25.0) | 17 | 1(5.9) | 2.042 | 0.153 | 76 | 23(30.3) | 33 | 1(3.0) | 9.938 | 0.002 |
| Laboratory examinations |  |  |  |  |  |  |  |  |  |  |  |  |
| WBC, × 10<sup>9</sup>/L, median (IQR) | 92 | 7.9(4.3,11.7) | 17 | 11.1(8.9,13.9) | -2.622 | 0.009 | 76 | 7.6(3.8,11.1) | 33 | 10.4(7.5,15.2) | -3.242 | 0.001 |
| CRP, mg/L, median (IQR) | 92 | 17.4(5.8,34.3) | 17 | 11.5(4.4,56.3) | -0.167 | 0.867 | 76 | 13.1(2.5,32.6) | 33 | 22.6(7.3,45.2) | -1.636 | 0.102 |
| PCT, ng/mL, median (IQR) | 73 | 0.3(0.2,0.5) | 9 | 0.2(0.1,10.5) | -0.334 | 0.739 | 60 | 0.3(0.2,0.5) | 22 | 0.4(0.2,0.8) | -1.136 | 0.256 |
| Plt, × 10<sup>9</sup>/L, median (IQR) | 92 | 226.5(109.0,306.0) | 17 | 414.0(266.0,565.5) | -4.021 | < 0.001 | 76 | 212.5(64.0,298.3) | 33 | 311.0(253.5,380.5) | -4.327 | < 0.001 |
| ALT, U/L, median (IQR) | 92 | 30.0(16.3,62.6) | 17 | 23.0(15.0,34.5) | -1.282 | 0.200 | 76 | 26.2(16.0,55.8) | 33 | 27.0(19.6,57.2) | -0.571 | 0.568 |
| AST, U/L, median (IQR) | 92 | 48.2(36.0,81.1) | 17 | 38.0(30.5,52.5) | -1.716 | 0.086 | 76 | 47.0(33.2,69.8) | 33 | 48.6(36.0,92.4) | -0.966 | 0.334 |
| Total bilirubin, umol/L, median (IQR) | 91 | 5.4(3.6,10.5) | 16 | 5.1(3.7,8.3) | -0.135 | 0.892 | 74 | 5.7(4.0,10.7) | 33 | 4.1(3.0,6.8) | -1.886 | 0.059 |
| Serum creatinine, umol/L, median (IQR) | 92 | 27.5(21.0,39.5) | 17 | 24.0(17.0,36.0) | -1.090 | 0.276 | 76 | 29.0(21.0,37.7) | 33 | 23.6(20.2,39.5) | -1.029 | 0.303 |
| Blood lactate, mmol/L, median (IQR) | 91 | 2.2(1.7,2.9) | 17 | 2.0(1.5,2.5) | -0.959 | 0.338 | 75 | 2.1(1.7,2.9) | 33 | 2.4(1.7,3.0) | -0.478 | 0.633 |
| Serogroup, n (%) |  |  |  |  |  |  |  |  |  |  |  |  |
| B |  | 24(26.1) |  | 4(24.5) | 0.000 | 1.000 |  | 14(18.4) |  | 14(42.4) | 6.945 | 0.008 |
| C | 92 | 26(28.3) | 17 | 8(47.1) | 2.362 | 0.124 | 76 | 25(32.9) | 33 | 9(27.3) | 0.339 | 0.560 |
| D |  | 27(29.3) |  | 2(11.8) | 1.461 | 0.227 |  | 25(32.9) |  | 4(12.1) | 5.085 | 0.024 |
| E |  | 8(8.7) |  | 0(0) | 0.573 | 0.449 |  | 5(6.6) |  | 3(9.1) | 0.004 | 0.950 |
| non A-E |  | 7(7.6) |  | 3(17.6) | 0.740 | 0.390 |  | 7(9.2) |  | 3(9.1) | 0.000 | 1.000 |
| Source of infection |  |  |  |  |  |  |  |  |  |  |  |  |
| BSI, n (%) | / | / | / | / | / | / |  | 66 |  | 26 | 1.134 | 0.287 |
| Non-BSI, n (%) | / | / | / | / | / | / |  | 10 |  | 7 |  |  |
| Clinical presentations, n (%) |  |  |  |  |  |  |  |  |  |  |  |  |
| Fever |  | 69(75.0) |  | 10(58.8) | 1.882 | 0.170 |  | 52(68.4) |  | 27(81.8) | 2.070 | 0.150 |
| Gastrointestinal symptoms |  | 35(38.0) |  | 4(23.5) | 1.315 | 0.251 |  | 20(26.3) |  | 19(57.6) | 9.785 | 0.002 |
| Cough |  | 7(7.6) |  | 2(11.8) | 0.327 | 0.567 |  | 6(7.9) |  | 3(9.1) | 0.043 | 0.835 |
| Hematological symptoms | 92 | 18(19.6) | 17 | 1(5.9) | 1.866 | 0.172 | 76 | 19(25.0) | 33 | 0(0) | 9.992 | 0.002 |
| Bone and joint |  | 4(4.3) |  | 9(52.9) | 32.255 | <<br>0.001 |  | 10(13.2) |  | 3(9.1) | 0.362 | 0.547 |
| Others |  | 7(7.6) |  | 2(11.8) | 0.009 | 0.926 |  | 6(7.9) |  | 3(9.1) | 0.000 | 1.000 |
| pSOFA≥2, n (%) | 92 | 50(54.3) | 17 | 5(29.4) | 3.569 | 0.059 | 76 | 34(44.7) | 33 | 20(60.6) | 2.318 | 0.128 |
| Outcomes |  |  |  |  |  |  |  |  |  |  |  |  |
| Duration of effective antibiotic use, days, median (IQR) | 92 | 9.0(7.0,12.0) | 17 | 25.0(11.5,42.0) | -4.091 | <<br>0.001 | 76 | 10.0(7.0,13.8) | 33 | 9.0(6.5,13.0) | -0.754 | 0.451 |
| Length of hospital stay, days, median (IQR) | 92 | 12.0(9.0,17.8) | 17 | 17.0(10.0,53.0) | -1.945 | 0.052 | 76 | 12.0(9.0,26.5) | 33 | 13.0(8.5,17.0) | -0.423 | 0.672 |
| Time to positive blood culture, hours, median (IQR) | 92 | 15.8(13.0,21.2) | 17 | 16.8(8.4,46.5) | -0.359 | 0.719 | 76 | 15.7(12.6,21.3) | 33 | 16.4(12.3,25.9) | -0.155 | 0.877 |
| Death, n (%) | 92 | 3(3.3) | 17 | 0(0) | 0.570 | 0.450 | 76 | 3(3.9) | 33 | 0(0) | 1.339 | 0.247 |

Compared to children over 1 year old, infants aged 1 year or less with iNTS infection had a higher proportion of serogroups B strains(42.4% vs. 18.4%, *P* = 0.008), a lower proportion of serogroups D strains (12.1% vs. 32.9%, *P* = 0.024), a higher prevalence of gastrointestinal symptoms (57.6% vs. 26.3%, *P* = 0.002), a lower proportion of hematological system involvement (0% vs. 25.0%, *P* = 0.002), a lower proportion of underlying diseases (21.2% vs. 40.8%, *P* = 0.049), a lower proportion of tumor (0% vs. 27.6%, *P* = 0.001), a lower proportion of deep venous catheterization (3.0% vs. 30.3%, *P* = 0.002). There were no significant differences in other parameters. (Tables 1)

In the strain serogroups, there were 28 cases (36.8%) in serogroup B, 34 cases (44.7%) in serogroup C, 29 cases (38.2%) in serogroup D, 8 cases in serogroup E, and 10 cases in non-A-E serogroup. The overall resistance rate of ampicillin was 53.6% (45/84), ceftriaxone resistance rate was 17.8% (19/107), SMZ resistance rate was 27.1% (29/107), ciprofloxacin insensitivity rate was 35.3% (30/85), chloramphenicol resistance rate was 35.6% (16/45), and carbapenem resistance rate was 0%. There were significant differences in the resistance of ampicillin, ceftriaxone, SMZ, and chloramphenicol among different NTS serogroups, while there was no significant difference in ciprofloxacin insensitivity rate. (Table 2)

**Table 2.** Antibiotic resistance of different Serogroups of NTS to six antibiotics.

| Antibiotic | Serogroup B | Serogroup C | Serogroup D | Serogroup E | Serogroup non A-E | $\chi^2$ | P |
| --- | --- | --- | --- | --- | --- | --- | --- |
| Ampicillin | 12/21(57.1%) | 9/29(31.0%) | 19/25(76.0%) | 3/3(100%) | 2/6(33.3%) | 16.222 | 0.003 |
| Ceftriaxone | 8/28(28.6%) | 2/33(6.1%) | 4/28(14.3%) | 5/8(62.5%) | 0/10(0%) | 17.941 | 0.001 |
| SMZ | 15/28(53.6%) | 6/33(18.2%) | 2/28(7.1%) | 5/8(62.5%) | 1/10(10.0%) | 23.573 | <0.001 |
| Ciprofloxacin | 11/21(52.4%) | 7/29(24.1%) | 9/26(34.6%) | 1/3(33.3%) | 2/6(33.3%) | 4.254 | 0.373 |
| Chloramphenicol | 9/11(81.8%) | 4/16(25.0%) | 2/15(13.3%) | 1/1(100%) | 0/2(0%) | 18.368 | 0.001 |
| Carbapenems | 0/20(0%) | 0/17(0%) | 0/17(0%) | 0/8(0%) | 0/7(0%) | / | / |

## Discussion

Among the 109 cases of iNTS, nearly 2/3 (65.1%) were boys, 38 cases had underlying diseases, and nearly half had acute lymphoblastic leukemia. The three death cases all came from bloodstream infection in children with hematological tumors after chemotherapy. The pSOFA score increased significantly. Due to the small number of death cases, statistical analysis of high-risk factors cannot be performed. pSOFA is helpful for disease assessment and prognosis judgment. It should be mentioned that the Psofa score indicators include serum creatinine and bilirubin, which limits early clinical application(5). Different from the total case fatality rate estimated in global reviews of 14.7%(3). there were few deaths in this study. Considering the different underlying disease spectrum, there were no children with HIV, which may also be the reason for the large difference in mortality. We noticed that there were no fatal cases reported in NTS bloodstream infection studies from Malaysia(6), Israel(2), Japan(7), and Taiwan, China(8).

The incidence of iNTS in children in both centers is related to temperature. During the high temperature season in summer, the number of incidences reaches a peak, while in winter and spring, the number of incidences decreases significantly. Possible factors include rising temperatures, easy reproduction of bacteria, easy deterioration of food, and water pollution, which are all risk factors for increasing NTS infection. Mohan et al. from Malaysia reported that there was no significant correlation between the monthly incidence of iNTS and the average monthly rainfall(9). In Hong Kong, China, it was reported that the peak of hospitalization for NTS infection was in June and the trough was in April(10). It shows that different latitudes in different regions are related to the seasonal characteristics of NTS incidence.

In this group of studies, the most common were BSI (84.4%) and bone and joint infection (12.8%). The clinical characteristics of the two groups and the antibiotic resistance are basically similar. The non-BSI iNTS group has a younger age of onset and longer antibiotic treatment time, suggesting that the non-BSI iNTS may have experienced a BSI process. Infections caused by blood circulation dissemination, it will take longer to clear the germs that spread the lesions. The higher white blood cell and platelet counts in the non-BSI group suggest a more severe inflammatory response in cases of non-BSI. Clinically, joint infection may spread hematogenously or directly spread from open wounds. This data found that the clinical parameters of the BSI group and the non-BSI iNTS group were mostly the same, and the clinical characteristics were similar, suggesting that the invasive infection group without BSI had bloodstream infection in early stage and spread the lesions through the blood circulation. The antibiotic treatment time in the non-BSI iNTS group was 2.5 times longer, indicating that it takes longer to clear bacteria from bone marrow, joints, cerebrospinal fluid and other lesions. Two studies in the United States compared the duration of intravenous antibiotic treatment for NTS BSI in children with >3 days to ≤3 days(11), and compared the duration of <7 days with ≥7 days(12). The clinical outcomes were good, and recommendations with oral antibiotics can be made as soon as possible. However, these two groups of cases were mild children with no invasive infection lesions and no underlying diseases, which were different from the cases studied in our groups.

Comparing the age groups, it can be found that serogroup B is more common in the infant group and serogroup D is less common. Clinical symptoms such as diarrhea and vomiting are more common. The proportion of older children with underlying diseases (mainly leukemia) and deep vein catheterization is higher than that of infants, indicating that with the age increases, acquired immune function strengthens, and the number of iNTS infections among children with normal immune function gradually decreases. The study by Sirinavin et al. from Thailand reported a group of 75 infant iNTS cases, including transient bacteremia (5), bacteremia without localized infection (37), bone and joint infection (5) and meningitis (28); while in our two centers, the cases were primarily bloodstream infections, only 2 cases of central nervous system infection, indicating a different spectrum of the disease. This also highlights the importance for clinicians to be vigilant about intracranial infections in infant iNTS infections(13).

NTS from different specimen sources are similar in antibiotic resistance, suggesting that infections at other invasion sites may also be caused by dissemination of BSI. In this study, the serogroups were predominantly B, C, and D, with relatively few serogroups E and other groups, which is similar to what was reported by Megged et al(2). In the NTS BSI reported by Lee et al. in Taiwan, serogroup D was the most common(14), their study showed that the ampicillin resistance rate was 53%, the third-generation cephalosporin resistance rate gradually increased to 11%, and the ciprofloxacin resistance rate dropped from 20% to 11%. In this study, ampicillin resistance exceeded 50%, carbapenem resistance was 0%, SMZ resistance was 27%, ceftriaxone resistance was 18%, and ciprofloxacin was mostly intermediate, with a proportion close to 35%. Our data indicate the emergence of extensive resistance to ampicillin, suggesting that Salmonella has extensively produced β-lactamase. Most are sensitive to ceftriaxone, but the resistance rate continues to rise. Some studies have used pulsed field gel electrophoresis to identify Salmonella strains that are resistant to ceftriaxone and identified the blaCMY-2 gene on the IncI1 plasmid; through horizontal gene transfer, the MIC of ceftriaxone for the recipient strain increased from 0.125 μg/mL to 32 μg/mL(15). The ciprofloxacin resistance rate has not declined because there are more intermediaries, which is closely related to the breakpoints defined by the Clinical Laboratory Standards Institute guidelines. Comparing the antibiotic resistance of different serogroups, it was found that serogroup E had the highest resistance rate to ampicillin, ceftriaxone, SMZ, and chloramphenicol. The resistance rate of ceftriaxone in serogroup C was 6.1%, while the resistance rate in serogroup E was the highest at 62.5%. To date, Ceftriaxone is still the first-line drug and serogroups has a certain guiding role in the selection of antibiotics. It is noteworthy that the drug resistance rate of Salmonella is gradually increasing. Garcia et al. in Peru reported multidrug-resistant Salmonella serovar Infantis (from serogroup C) were only sensitive to carbapenems(16). In our study, all carbapenems are sensitive. Considering that children with iNTS are more likely to have underlying diseases and have immune dysfunction, carbapenem antibiotics can be used as second-line antibiotics when severe invasive infections occur.

This study is a retrospective study, and the strains isolated from the specimens are only grouped into serogroups without serotype classification, which limits further analysis of molecular characteristics. Secondly, the specimens of invasive infection are mainly from blood, bone and joint, and there is very little from CSF and pleural effusion, which limits in-depth comparative analysis of clinical characteristics. However, the 109 iNTS cases are still a relatively large cohort and can provide meaningful reference for subsequent research on the clinical characteristics and treatment of Salmonella.

Conclusion: The clinical manifestations and treatment of iNTS infection in children are related to the site of invasion. Ceftriaxone is still the first-line antibiotic, but the resistance is gradually increasing. Carbapenems can be considered as second-line antibiotics in severe cases.

## Data Availability

All relevant data are within the manuscript and its Supporting Information files.

